# Characteristics and Factors Associated with COVID-19 Infection, Hospitalization, and Mortality Across Race and Ethnicity

**DOI:** 10.1101/2020.10.14.20212803

**Authors:** Chengzhen L. Dai, Sergey A. Kornilov, Ryan T. Roper, Hannah Cohen-Cline, Kathleen Jade, Brett Smith, James R. Heath, George Diaz, Jason D. Goldman, Andrew T. Magis, Jennifer J. Hadlock

## Abstract

**Background:** Data on the characteristics of COVID-19 patients disaggregated by race/ethnicity remain limited. We evaluated the sociodemographic and clinical characteristics of patients across racial/ethnic groups and assessed their associations with COVID-19 outcomes.

**Methods:** This retrospective cohort study examined 629,953 patients tested for SARS-CoV-2 in a large health system spanning California, Oregon, and Washington between March 1 and December 31, 2020. Sociodemographic and clinical characteristics were obtained from electronic health records. Odds of SARS-CoV-2 infection, COVID-19 hospitalization, and in-hospital death were assessed with multivariate logistic regression.

**Results:** 570,298 patients with known race/ethnicity were tested for SARS-CoV-2, of whom 27.8% were non-White minorities. 54,645 individuals tested positive, with minorities representing 50.1%. Hispanics represented 34.3% of infections but only 13.4% of tests. While generally younger than White patients, Hispanics had higher rates of diabetes but fewer other comorbidities. 8,536 patients were hospitalized and 1,246 died, of whom 56.1% and 54.4% were non-White, respectively. Racial/ethnic distributions of outcomes across the health system tracked with state-level statistics. Increased odds of testing positive and hospitalization were associated with all minority races/ethnicities. Hispanic patients also exhibited increased morbidity, and Hispanic race/ethnicity was associated with in-hospital mortality (OR: 1.39 [95% CI: 1.14-1.70]).

**Conclusion:** Major healthcare disparities were evident, especially among Hispanics who tested positive at a higher rate, required excess hospitalization and mechanical ventilation, and had higher odds of in-hospital mortality despite younger age. Targeted, culturally-responsive interventions and equitable vaccine development and distribution are needed to address the increased risk of poorer COVID-19 outcomes among minority populations.

**Key points:** Racial/ethnic disparities are evident in the disaggregated characteristics of COVID-19 patients. Minority patients experience increased odds of SARS-CoV-2 infection and COVID-19 hospitalization. Hospitalized Hispanic patients presented with more severe illness, experienced increased morbidity, and faced increased mortality.

## Introduction

Since the coronavirus disease 2019 (COVID-19) was first reported in Washington state, the United States has documented the highest number of confirmed cases and deaths in the world. Increasing evidence have indicated that COVID-19 disproportionately affects patients of minority race and ethnicity.[1–3] While reports have identified different rates of infection, hospitalization, and mortality among minority populations, there is limited information on the characteristics of COVID-19 patients disaggregated by race/ethnicity.[4,5]

The prevalence of comorbidities and social environments vary between racial/ethnic groups.[6,7] Some of these characteristics, including obesity and crowded housing, are potential risk factors for COVID-19 and disease severity.[8,9] Therefore, understanding how the characteristics of patients differ between races/ethnicities and which factors are associated with disease outcomes are critical for public health and designing community-based interventions. Unfortunately, such detailed characteristics remain sparse and certain racial/ethnic groups, specifically Asian Americans, Native Hawaiians/Pacific Islanders (NH/PI), and American Indians/Alaska Natives (AI/AN) remain yet to be characterized in detail.[10,11] Furthermore, while sociodemographic and health characteristics vary across geography, multi-state comparisons are limited. The objective of this study therefore is to examine the characteristics of and factors associated with SARS-CoV-2 infection, hospitalization with COVID-19, and in-hospital mortality in a large diverse population of patients in a large health system operating in California, Oregon, and Washington.

## Method

### Study Design, Setting, and Population

This retrospective cohort study included patients from California, Oregon, and Washington who were tested for SARS-CoV-2 with a polymerase chain reaction (PCR) assay of a nasopharyngeal sample and were seen at a Providence St. Joseph Heath (PSJH) facility between March 1, 2020 and December 31, 2020. In-hospital outcomes were monitored through January 31, 2020. PSJH is one of the largest health systems in the US. In 2019, approximately 3.5 million patients in California, Oregon, and Washington received care at the facilities included in this study, of whom 62.5% identified as non-Hispanic White; 11.5% as Hispanic; 6.8% as non-Hispanic Asian American; 3.6 % as non-Hispanic Black; 0.6% non-Hispanic American Indian/Alaska Native; 0.5% non-Hispanic Native Hawaiian/Pacific Islander; 5.7% non-Hispanic Other; and 8.8% Unknown. The protocol for this study was approved by the PSJH Institutional Review Board (IRB#: STUDY2020000203).

### Data Collection

Patient demographic and clinical data were extracted from PSJH’s Epic electronic health record system. Patients with a positive PCR test for SARS-CoV-2 were considered to have a confirmed SARS-CoV-2 infection. For patients who had multiple tests, only the initial positive test result was considered. Extracted demographic data included age, sex, race, ethnicity, and insurance plan. 85 patients with missing sex were excluded from the study. Missing race or ethnicity were grouped as Unknown. ZIP codes were used to identify the neighborhood-level median income, crowded housing (>1 person per room), minority population (race/ethnicity except non-Hispanic white), and limited English-proficient speakers from the US Census Bureau’s American Community Survey. Clinical data include underlying medical conditions identified using ICD-10-CM codes or direct clinical measurements linked to past encounters between January 1, 2019 and the date of SARS-CoV-2 testing. We included underlying medical conditions that have previously been associated with COVID-19.[12,13] Charlson Comorbidity Index (CCI) was used to capture the risk from multiple comorbidities. Obesity and hypertension, which are not part of CCI, were also included. We used previously-defined diagnosis codes for components of CCI;[14] I10 for hypertension and BMI classified based on Center for Disease Control definitions for obesity. Inpatient encounter data included presenting vital signs, baseline laboratories, supplemental oxygen use, acute diagnoses (COVID-19: U07.1; lower respiratory infection: J22; acute respiratory distress syndrome: J80; respiratory failure: J96; pneumonia: J12.89), length of stay, transfer to intensive care unit, and discharge disposition.

For comparison with state-level data, COVID-19 cases and deaths for California, Oregon, and Washington was obtained from the COVID Racial Data Tracker, which aggregates historical data from state agencies.[15] Data on COVID-19 hospitalization for California and Oregon was obtained from the Center for Disease Control’s COVID-NET while Washington data was obtained from the COVID Racial Data Tracker.[15,16]

### Statistical Analysis

We compared the sociodemographic and clinical characteristics of patients across COVID-19 outcomes and race/ethnicity categories, defined as Hispanic; non-Hispanic Black (Black); non-Hispanic Asian American (Asian); non-Hispanic Native Hawaiian and Pacific Islander (NH/PI); non-Hispanic American Indian and Alaska Native (AI/AN); non-Hispanic white (White); and non-Hispanic other (Other), which includes multi-race/ethnicity. For COVID-19 hospitalized patients, presenting clinical characteristics were available and included clinical status on the World Health Organization (WHO) 9-point Clinical Progression Scale;[17] presenting vitals within the first 6 hours of admissions; and baseline laboratory test results within 24 hours of admissions. The WHO Clinical Progression Scale was developed to measure clinical illness of an COVID-19 infection and consists of the following categories: 0–uninfected; 1–ambulatory, no limitation of activity; 2–ambulatory, limitation of activity; 3–hospitalized, no oxygen therapy; 4–hospitalized, oxygen by mask or nasal prongs; 5–hospitalized, non-invasive ventilation or high-flow oxygen; 6–hospitalized, intubation and mechanical ventilation; 7–hospitalized, ventilation + additional organ support; 8–death.

Associations with SARS-CoV-2 infection, COVID-19 hospitalization, and in-hospital mortality was assessed using mixed-effect logistic regression models with state and month of diagnosis nested random effect variables to account for geographic and temporal variations. For each outcome, we fitted both unadjusted univariate models and adjusted multivariate models. All multivariate models included race/ethnicity as an independent variable, with demographic factors (age; age-squared; sex), socioeconomic factors (insurance; neighborhood median income, crowded housing, limited English proficiency, and minority), and comorbidities (CCI; hypertension; obesity) as covariates. An age-squared term was included in addition to age to capture the non-linear relationship between COVID-19 outcomes and age.[18] For analyses of hospital mortality, additional covariates included presenting WHO Clinical Progression Scale score and baseline lab results. Covariates were selected based on previously identified risk factors and patterns of missingness and collinearity. Certain characteristics—BMI, insurance coverage, and baseline lab results—were not available for all patients. Alanine transaminase was excluded due to high correlation with aspartate transaminase and had higher missingness. For variables with less than 20% missingness, missing values were imputed with multiple imputation by fully conditional specification (15 imputations). Statistical analyses were performed with R v3.6.2.

## Results

### Characteristics of Patients Tested for SARS-CoV-2

A total of 629,953 patients tested for SARS-CoV-2 were included. 570,298 patients (90.5%) reported race/ethnicity, of which 72.2% were White, 13.4% were Hispanic, 5.4% were Asian, 3.8% Black, 0.9% were AI/AN, 0.6% were NH/PI, and 3.7% were Other (**Table 1**). The mean age among all tested patients was 51.5±19.4 years and 57.0% were female. The majority of patients (53.7%) had public insurance (Medicaid: 30.0%; Medicare: 23.7%), with higher percentages of Hispanic, Black, and AI/AN patients on Medicaid than White, Asian, and NH/PI patients (**Table S1**). The most common comorbidities were obesity (37.1%), hypertension (23.3%), diabetes (9.4%) and asthma (6.5%). The median score on the Charlson Comorbidity Index (CCI) was 1.0 (95% CI: 0.0-3.0).

**Table 1.** Characteristics of patients tested and positive for SARS-CoV-2 and hospitalized for COVID-19.

|  | Tested patients<br>No. (%) | Positive patients<br>No. (%) | Hospitalized patients<br>No. (%) |
| --- | --- | --- | --- |
| **Socio-demographics** |  |  |  |
| Total no. of patients | 629,953 | 54,645 | 8,536 |
| Age (years) | 51.5 ± 19.4 | 47.8 ± 19.2 | 64.7 ± 17.6 |
| Sex |  |  |  |
| Female | 359,246 (57.0%) | 28,765 (52.6%) | 3,885 (45.5%) |
| Male | 270,707 (43.0%) | 25,880 (47.4%) | 4,651 (54.5%) |
| Known race/ethnicity | 570,298 (90.5%) | 49,081 (89.8%) | 8,210 (96.2%) |
| Hispanic | 76,300 (13.4%) | 16,836 (34.3%) | 2,898 (35.3%) |
| Black | 21,709 (3.8%) | 2,066 (4.2%) | 368 (4.5%) |
| Asian | 30,736 (5.4%) | 2,456 (5.0%) | 533 (6.5%) |
| NH/PI | 3,460 (0.6%) | 468 (1.0%) | 98 (1.2%) |
| AI/AN | 5,201 (0.9%) | 434 (0.9%) | 77 (0.9%) |
| White | 411,852 (72.2%) | 24,468 (49.9%) | 3,751 (45.7%) |
| Other | 21,040 (3.7%) | 2,353 (4.8%) | 485 (5.9%) |
| Unknown race/ethnicity | 59,655 (9.5%) | 5,564 (10.2%) | 326 (3.8%) |
| Insurance |  |  |  |
| Commercial | 280,300/616,048 (45.5%) | 23,617/52,764 (44.8%) | 1,494/8,432 (17.7%) |
| Medicaid | 184,547/616,048 (30.0%) | 20,937/52,764 (39.7%) | 4,327/8,432 (51.3%) |
| Medicare | 146,041/616,048 (23.7%) | 6,989/52,764 (13.2%) | 2,495/8,432 (29.6%) |
| Uninsured/Self-pay | 5,001/616,048 (0.8%) | 1,211/52,764 (2.3%) | 113/8,432 (1.3%) |
| Other insurance | 159/616,048 (0.0%) | 10/52,764 (0.0%) | 3/8,432 (0.0%) |
| Neighborhood-level |  |  |  |
| Median income | 74,604 ± 25,144 | 68,522 ± 21,589 | 68,843 ± 22,432 |
| % Crowded housing | 4.4 ± 4.6 | 6.4 ± 6.4 | 7.8 ± 7.1 |
| % Minority | 34.3 ± 21.4 | 42.4 ± 26.3 | 49.4 ± 27.0 |
| % Limited English | 9.3 ± 8.6 | 12.8 ± 10.5 | 15.9 ± 11.3 |
| **Comorbidities** |  |  |  |
| Hypertension | 146,957 (23.3%) | 10,847 (19.8%) | 3,433 (40.2%) |
| Diabetes | 59,452 (9.4%) | 5,946 (10.9%) | 2,419 (28.3%) |
| Weight |  |  |  |
| Underweight | 11,719/548,795 (2.1%) | 781/47,184 (1.7%) | 262/8,503 (3.1%) |
| Normal | 159,034/548,795 (29.0%) | 11,277/47,184 (23.9%) | 2,070/8,503 (24.3%) |
| Overweight | 174,105/548,795 (31.7%) | 15,363/47,184 (32.6%) | 2,573/8,503 (30.3%) |
| Class 1 Obesity | 108,833/548,795 (19.8%) | 10,499/47,184 (22.3%) | 1,801/8,503 (21.2%) |
| Class 2 Obesity | 52,914/548,795 (9.6%) | 5,214/47,184 (11.1%) | 924/8,503 (10.9%) |
| Class 3 Obesity | 42,190/548,795 (7.7%) | 4,050/47,184 (8.6%) | 873/8,503 (10.3%) |
| Chronic respiratory disease |  |  |  |
| Asthma | 40,808 (6.5%) | 2,894 (5.3%) | 574 (6.7%) |
| COPD | 25,313 (4.0%) | 1,424 (2.6%) | 712 (8.3%) |
| Cardiovascular disease |  |  |  |
| Coronary artery disease | 34,549 (5.5%) | 1,976 (3.6%) | 829 (9.7%) |
| Myocardial Infarction | 13,979 (2.2%) | 872 (1.6%) | 473 (5.5%) |
| Congestive heart failure | 33,675 (5.3%) | 2,128 (3.9%) | 1,129 (13.2%) |
| Kidney disease | 35,154 (5.6%) | 2,897 (5.3%) | 1,469 (17.2%) |
| Liver disease | 19,772 (3.1%) | 1,353 (2.5%) | 345 (4.0%) |
| Cancer | 38,453 (6.1%) | 1,663 (3.0%) | 539 (6.3%) |
| Charlson Comorbidity Index | 1.0 (0.0 - 3.0) | 0.0 (0.0 - 2.0) | 3.0 (2.0 - 6.0) |
| **Geographic distribution** |  |  |  |
| California | 157,407 (25.0%) | 18,701 (34.2%) | 4,237 (49.6%) |
| Oregon | 167,874 (26.6%) | 13,486 (24.7%) | 1,047 (12.3%) |
| Washington | 304,672 (48.4%) | 22,458 (41.1%) | 3,252 (38.1%) |

### Characteristics of Patients Positive for SARS-CoV-2 Infection

54,645 patients (8.7%) of the 629,953 patients tested for SARS-CoV-2 were positive. Among the 49,081 patients with known race/ethnicity, the rate of positive test results was higher for minority patients than White patients (5.9%; **Table S1**). Hispanic and NH/PI patients had the highest rates (22.1% and 13.5%, respectively). Consequently, the racial/ethnic composition of SARS-CoV-2 infected patients were 49.9% White, 34.3% Hispanic, 5.0% Asian, 4.8% Other, 4.2% Black, 1.0% NH/PI (1.4%), and 0.9% AI/AN patients (**Table 1**).

Among all SARS-CoV-2 infected patients, the mean age was 47.8±19.2 years and 52.6% were female (**Table 1**). Compared to White patients, mean ages were lower among patients of minority race/ethnicity, except for Asians, which had a similar mean age (**Table 2**). CCI scores were also lower among minority patients. The prevalence of diabetes, however, was higher among minority patients. Additionally, relative to White patients, Hispanic, Black, NH/PI, and AI/AN patients had higher prevalence of obesity; Asian, Black, NH/PI, and AI/AN patients had higher prevalence of hypertension; and Black, NH/PI, and AI/AN patients had higher prevalence of both asthma and kidney disease.

**Table 2.** Characteristics of SARS-CoV-2 infected patients by race/ethnicity.

|  | Hispanic<br>No. (%) | Black<br>No. (%) | Asian<br>No. (%) | NH/PI<br>No. (%) | AI/AN<br>No. (%) | White<br>No. (%) | Other<br>No. (%) | Unknown<br>No. (%) |
| --- | --- | --- | --- | --- | --- | --- | --- | --- |
| **Socio-demographics** |  |  |  |  |  |  |  |  |
| Total no. of patients | 16,836 | 2,066 | 2,456 | 468 | 434 | 24,468 | 2,353 | 5,564 |
| Hospitalized | 2,898 (17.2%) | 368 (17.8%) | 533 (21.7%) | 98 (20.9%) | 77 (17.7%) | 3,751 (15.3%) | 485 (20.6%) | 326 (5.9%) |
| Age (years) | 44.7 ± 17.3 | 46.4 ± 18.5 | 51.7 ± 19.4 | 45.6 ± 16.5 | 44.8 ± 17.9 | 51.1 ± 20.3 | 49.7 ± 19.1 | 41.8 ± 17.4 |
| Sex |  |  |  |  |  |  |  |  |
| Female | 9,064 (53.8%) | 1,080 (52.3%) | 1,347 (54.8%) | 262 (56.0%) | 252 (58.1%) | 12,842 (52.5%) | 1,181 (50.2%) | 2,737 (49.2%) |
| Male | 7,772 (46.2%) | 986 (47.7%) | 1,109 (45.2%) | 206 (44.0%) | 182 (41.9%) | 11,626 (47.5%) | 1,172 (49.8%) | 2,827 (50.8%) |
| Insurance |  |  |  |  |  |  |  |  |
| Commercial | 5,878/16,192 (36.3%) | 688/2,015 (34.1%) | 1,226/2,401 (51.1%) | 195/457 (42.7%) | 113/424 (26.7%) | 11,513/24,077 (47.8%) | 904/2,271 (39.8%) | 3,100/4,927 (62.9%) |
| Medicaid | 8,751/16,192 (54.0%) | 1,119/2,015 (55.5%) | 861/2,401 (35.9%) | 204/457 (44.6%) | 260/424 (61.3%) | 7,464/24,077 (31.0%) | 1,108/2,271 (48.8%) | 1,170/4,927 (23.7%) |
| Medicare | 863/16,192 (5.3%) | 168/2,015 (8.3%) | 290/2,401 (12.1%) | 41/457 (9.0%) | 51/424 (12.0%) | 4,964/24,077 (20.6%) | 214/2,271 (9.4%) | 398/4,927 (8.1%) |
| Uninsured/Self-pay | 693/16,192 (4.3%) | 40/2,015 (2.0%) | 23/2,401 (1.0%) | 17/457 (3.7%) | 0 (0.0%) | 135/24,077 (0.6%) | 44/2,271 (1.9%) | 259/4,927 (5.3%) |
| Other insurance | 7/16,192 (0.0%) | 0 (0.0%) | 1/2,401 (0.0%) | 0 (0.0%) | 0 (0.0%) | 1/24,077 (0.0%) | 1/2,271 (0.0%) | 0 (0.0%) |
| Neighborhood-level |  |  |  |  |  |  |  |  |
| Median income | 63,454 ± 17,110 | 65,525 ± 19,805 | 74,491 ± 23,569 | 65,528 ± 20,276 | 68,286 ± 18,073 | 70,511 ± 22,886 | 71,427 ± 23,518 | 72,630 ± 23,845 |
| % Crowded housing | 10.3 ± 7.8 | 6.9 ± 6.0 | 5.7 ± 4.8 | 5.2 ± 4.8 | 3.6 ± 2.7 | 3.7 ± 3.7 | 6.2 ± 5.4 | 6.5 ± 6.7 |
| % Minority | 59.0 ± 27.2 | 53.0 ± 26.7 | 45.8 ± 21.0 | 39.0 ± 22.0 | 29.5 ± 15.9 | 29.2 ± 18.3 | 44.7 ± 22.5 | 44.7 ± 26.1 |
| % Limited English | 19.2 ± 11.0 | 14.5 ± 9.3 | 14.0 ± 8.7 | 11.1 ± 8.4 | 6.2 ± 5.2 | 8.0 ± 7.7 | 14.4 ± 9.8 | 12.9 ± 10.5 |
| **Comorbidities** |  |  |  |  |  |  |  |  |
| Hypertension | 2,726 (16.2%) | 585 (28.3%) | 620 (25.2%) | 117 (25.0%) | 113 (26.0%) | 5,718 (23.4%) | 464 (19.7%) | 504 (9.1%) |
| Diabetes | 2,025 (12.0%) | 338 (16.4%) | 387 (15.8%) | 105 (22.4%) | 63 (14.5%) | 2,496 (10.2%) | 267 (11.3%) | 265 (4.8%) |
| BMI |  |  |  |  |  |  |  |  |
| Underweight | 147/15,062 (1.0%) | 40/1,832 (2.2%) | 88/2,162 (4.1%) | 6/423 (1.4%) | 6/391 (1.5%) | 401/21,621 (1.9%) | 37/2,088 (1.8%) | 56/3,605 (1.6%) |
| Normal | 2,519/15,062 (16.7%) | 416/1,832 (22.7%) | 976/2,162 (45.1%) | 81/423 (19.1%) | 70/391 (17.9%) | 5,713/21,621 (26.4%) | 527/2,088 (25.2%) | 975/3,605 (27.0%) |
| Overweight | 5,132/15,062 (34.1%) | 557/1,832 (30.4%) | 722/2,162 (33.4%) | 112/423 (26.5%) | 98/391 (25.1%) | 6,780/21,621 (31.4%) | 706/2,088 (33.8%) | 1,256/3,605 (34.8%) |
| Class 1 Obesity | 4,002/15,062 (26.6%) | 401/1,832 (21.9%) | 263/2,162 (12.2%) | 97/423 (22.9%) | 97/391 (24.8%) | 4,462/21,621 (20.6%) | 470/2,088 (22.5%) | 707/3,605 (19.6%) |
| Class 2 Obesity | 1,887/15,062 (12.5%) | 215/1,832 (11.7%) | 70/2,162 (3.2%) | 58/423 (13.7%) | 56/391 (14.3%) | 2,346/21,621 (10.9%) | 209/2,088 (10.0%) | 373/3,605 (10.3%) |
| Class 3 Obesity | 1,375/15,062 (9.1%) | 203/1,832 (11.1%) | 43/2,162 (2.0%) | 69/423 (16.3%) | 64/391 (16.4%) | 1,919/21,621 (8.9%) | 139/2,088 (6.7%) | 238/3,605 (6.6%) |
| Chronic respiratory disease |  |  |  |  |  |  |  |  |
| Asthma | 731 (4.3%) | 164 (7.9%) | 135 (5.5%) | 30 (6.4%) | 63 (14.5%) | 1,506 (6.2%) | 104 (4.4%) | 161 (2.9%) |
| COPD | 133 (0.8%) | 71 (3.4%) | 40 (1.6%) | 7 (1.5%) | 28 (6.5%) | 1,056 (4.3%) | 52 (2.2%) | 37 (0.7%) |
| Cardiovascular disease |  |  |  |  |  |  |  |  |
| Coronary artery disease | 281 (1.7%) | 54 (2.6%) | 97 (3.9%) | 26 (5.6%) | 17 (3.9%) | 1,346 (5.5%) | 86 (3.7%) | 69 (1.2%) |
| Myocardial Infarction | 170 (1.0%) | 46 (2.2%) | 52 (2.1%) | 14 (3.0%) | 9 (2.1%) | 495 (2.0%) | 45 (1.9%) | 41 (0.7%) |
| Congestive heart failure | 374 (2.2%) | 111 (5.4%) | 90 (3.7%) | 22 (4.7%) | 25 (5.8%) | 1,328 (5.4%) | 102 (4.3%) | 76 (1.4%) |
| Kidney disease | 755 (4.5%) | 170 (8.2%) | 159 (6.5%) | 46 (9.8%) | 29 (6.7%) | 1,530 (6.3%) | 117 (5.0%) | 91 (1.6%) |
| Liver disease | 474 (2.8%) | 45 (2.2%) | 93 (3.8%) | 13 (2.8%) | 27 (6.2%) | 592 (2.4%) | 50 (2.1%) | 59 (1.1%) |
| Cancer | 323 (1.9%) | 60 (2.9%) | 77 (3.1%) | 7 (1.5%) | 16 (3.7%) | 1,069 (4.4%) | 44 (1.9%) | 67 (1.2%) |
| Charlson Comorbidity Index | 0.0 (0.0 - 2.0) | 1.0 (0.0 - 3.0) | 1.0 (0.0 - 3.0) | 0.0 (0.0 - 2.0) | 1.0 (0.0 - 3.0) | 1.0 (0.0 - 3.0) | 1.0 (0.0 - 3.0) | 0.0 (0.0 - 1.0) |
| **Location of Testing** |  |  |  |  |  |  |  |  |
| Emergency | 5,114 (30.4%) | 652 (31.6%) | 422 (17.2%) | 117 (25.0%) | 70 (16.1%) | 3,367 (13.8%) | 663 (28.2%) | 371 (6.7%) |
| Inpatient | 2,854 (17.0%) | 385 (18.6%) | 538 (21.9%) | 98 (20.9%) | 73 (16.8%) | 3,994 (16.3%) | 503 (21.4%) | 343 (6.2%) |
| Urgent care | 4,142 (24.6%) | 390 (18.9%) | 628 (25.6%) | 106 (22.6%) | 70 (16.1%) | 8,029 (32.8%) | 538 (22.9%) | 1,322 (23.8%) |
| Other outpatient | 4,165 (24.7%) | 564 (27.3%) | 792 (32.2%) | 124 (26.5%) | 204 (47.0%) | 7,723 (31.6%) | 566 (24.1%) | 3,167 (56.9%) |
| Other service area | 561 (3.3%) | 74 (3.6%) | 75 (3.1%) | 23 (4.9%) | 17 (3.9%) | 1,355 (5.5%) | 83 (3.5%) | 361 (6.5%) |

Hispanic, Black, NH/PI, and AI/NH patients were more likely to have Medicaid insurance than White and Asian patients (**Table 2**). All minority patients were more likely than White patients to reside in neighborhoods with higher percentages of crowded housing and minorities. Hispanic, Asian, and Black patients in particular were also more likely to live in neighborhoods with a higher percentage of limited English proficient population. All minority patients were more likely to have received their tests in the emergency department than White patients.

### Characteristics of Patients Hospitalized with COVID-19

15.6% (n=8,536) of the patients who tested positive for COVID-19 were hospitalized. 8,210 patients (96.2%) had known race/ethnicity, of which 45.7% were White, 35.3% were Hispanic, 6.5% were Asian, 4.5% were Black, 1.2% were NH/PI, 0.9% were AI/AN, and 5.9% were Other (**Table 1**). The mean age of all patients was 64.7±17.6 years and 54.5% were male. The median CCI score was 3.0 (2.0-6.0) and the most common comorbidities were obesity (42.4%), hypertension (40.2%), and diabetes (28.3%).

White patients had the highest mean age (70.5±16.2 years; **Table 3**) and median CCI score (4.0 [3.0-7.0]). NI/PI patients had the lowest mean age (55.9±17.3 years), and lowest median CCI score (2.0 [1.0-5.0]). The prevalence of obesity was highest among AI/AN, NH/PI, and Hispanic patients and lowest among Asian patients. However, Hispanics had the lowest prevalence of hypertension while Asian and Black patients had the highest. The majority of Hispanic, Black, and AI/AN patients were on Medicaid; and Hispanic, Black and Asian patients resided in neighborhoods with higher percentages of crowded housing, minorities, and limited English speakers than other racial/ethnic subgroups.

**Table 3.** Characteristics of patients hospitalized with COVID-19 by race/ethnicity.

|  | Hispanic<br>No. (%) | Black<br>No. (%) | Asian<br>No. (%) | NH/PI<br>No. (%) | AI/AN<br>No. (%) | White<br>No. (%) | Other<br>No. (%) | Unknown<br>No. (%) |
| --- | --- | --- | --- | --- | --- | --- | --- | --- |
| **Demographic characteristics** |  |  |  |  |  |  |  |  |
| Total no. of patients | 2,898 | 368 | 533 | 98 | 77 | 3,751 | 485 | 326 |
| Age (years) | 57.6 ± 17.0 | 61.9 ± 16.7 | 67.4 ± 17.0 | 55.9 ± 17.3 | 58.3 ± 15.6 | 70.5 ± 16.2 | 65.5 ± 16.6 | 62.5 ± 17.2 |
| Sex |  |  |  |  |  |  |  |  |
| Female | 1,255 (43.3%) | 163 (44.3%) | 254 (47.7%) | 50 (51.0%) | 33 (42.9%) | 1,762 (47.0%) | 229 (47.2%) | 139 (42.6%) |
| Male | 1,643 (56.7%) | 205 (55.7%) | 279 (52.3%) | 48 (49.0%) | 44 (57.1%) | 1,989 (53.0%) | 256 (52.8%) | 187 (57.4%) |
| Insurance |  |  |  |  |  |  |  |  |
| Commercial | 546 (18.8%) | 66 (17.9%) | 141 (26.5%) | 29 (29.6%) | 10 (13.0%) | 532 (14.2%) | 99 (20.4%) | 71 (21.8%) |
| Medicaid | 1,873 (64.6%) | 229 (62.2%) | 250 (46.9%) | 46 (46.9%) | 46 (59.7%) | 1,461 (38.9%) | 264 (54.4%) | 158 (48.5%) |
| Medicare | 353 (12.2%) | 69 (18.8%) | 130 (24.4%) | 19 (19.4%) | 21 (27.3%) | 1,711 (45.6%) | 107 (22.1%) | 85 (26.1%) |
| Uninsured/Self-pay | 77 (2.7%) | 4 (1.1%) | 6 (1.1%) | 3 (3.1%) | 0 (0.0%) | 14 (0.4%) | 3 (0.6%) | 6 (1.8%) |
| Other insurance | 2 (0.1%) | 0 (0.0%) | 1 (0.2%) | 0 (0.0%) | 0 (0.0%) | 0 (0.0%) | 0 (0.0%) | 0 (0.0%) |
| Neighborhood-level |  |  |  |  |  |  |  |  |
| Median income | 62,443 ± 16,270 | 65,898 ± 21,196 | 76,028 ± 24,288 | 64,616 ± 20,996 | 63,676 ± 18,927 | 72,623 ± 24,352 | 72,011 ± 25,977 | 71,588 ± 25,702 |
| % Crowded housing | 12.4 ± 7.9 | 8.4 ± 6.9 | 6.6 ± 5.6 | 5.1 ± 4.5 | 3.9 ± 3.5 | 4.7 ± 4.6 | 7.5 ± 6.2 | 5.9 ± 5.6 |
| % Minority | 67.5 ± 24.8 | 61.8 ± 27.5 | 51.2 ± 21.5 | 40.0 ± 23.4 | 34.1 ± 20.4 | 35.0 ± 20.5 | 50.5 ± 23.2 | 42.7 ± 22.5 |
| % Limited English | 22.9 ± 10.7 | 16.6 ± 9.5 | 16.1 ± 9.5 | 11.4 ± 8.5 | 6.7 ± 6.1 | 10.7 ± 9.4 | 17.6 ± 10.8 | 12.9 ± 9.4 |
| **Comorbidities** |  |  |  |  |  |  |  |  |
| Hypertension | 861 (29.7%) | 191 (51.9%) | 242 (45.4%) | 43 (43.9%) | 35 (45.5%) | 1,751 (46.7%) | 189 (39.0%) | 121 (37.1%) |
| Diabetes | 851/2,893 (29.4%) | 145 (39.4%) | 182 (34.1%) | 43 (43.9%) | 24 (31.2%) | 966/3,744 (25.8%) | 123/484 (25.4%) | 85 (26.1%) |
| BMI |  |  |  |  |  |  |  |  |
| Underweight | 47/2,891 (1.6%) | 14/367 (3.8%) | 37/530 (7.0%) | 3 (3.1%) | 2 (2.6%) | 142/3,732 (3.8%) | 8/482 (1.7%) | 9 (2.8%) |
| Normal | 528/2,891 (18.3%) | 85/367 (23.2%) | 247/530 (46.6%) | 19 (19.4%) | 15 (19.5%) | 982/3,732 (26.3%) | 116/482 (24.1%) | 78 (23.9%) |
| Overweight | 919/2,891 (31.8%) | 105/367 (28.6%) | 165/530 (31.1%) | 25 (25.5%) | 20 (26.0%) | 1,069/3,732 (28.6%) | 161/482 (33.4%) | 109 (33.4%) |
| Class 1 Obesity | 735/2,891 (25.4%) | 77/367 (21.0%) | 52/530 (9.8%) | 20 (20.4%) | 17 (22.1%) | 737/3,732 (19.7%) | 109/482 (22.6%) | 54 (16.6%) |
| Class 2 Obesity | 348/2,891 (12.0%) | 40/367 (10.9%) | 14/530 (2.6%) | 12 (12.2%) | 10 (13.0%) | 399/3,732 (10.7%) | 51/482 (10.6%) | 50 (15.3%) |
| Class 3 Obesity | 314/2,891 (10.9%) | 46/367 (12.5%) | 15/530 (2.8%) | 19 (19.4%) | 13 (16.9%) | 403/3,732 (10.8%) | 37/482 (7.7%) | 26 (8.0%) |
| Chronic respiratory disease |  |  |  |  |  |  |  |  |
| Asthma | 131 (4.5%) | 32 (8.7%) | 38 (7.1%) | 13 (13.3%) | 13 (16.9%) | 297 (7.9%) | 31 (6.4%) | 19 (5.8%) |
| COPD | 56 (1.9%) | 35 (9.5%) | 26 (4.9%) | 3 (3.1%) | 13 (16.9%) | 530 (14.1%) | 33 (6.8%) | 16 (4.9%) |
| Cardiovascular disease |  |  |  |  |  |  |  |  |
| Coronary artery disease | 121 (4.2%) | 25 (6.8%) | 54 (10.1%) | 10 (10.2%) | 7 (9.1%) | 540 (14.4%) | 47 (9.7%) | 25 (7.7%) |
| Myocardial infarction | 96 (3.3%) | 28 (7.6%) | 35 (6.6%) | 6 (6.1%) | 4 (5.2%) | 245 (6.5%) | 33 (6.8%) | 26 (8.0%) |
| Congestive heart failure | 207 (7.1%) | 60 (16.3%) | 57 (10.7%) | 12 (12.2%) | 12 (15.6%) | 683 (18.2%) | 61 (12.6%) | 37 (11.3%) |
| Kidney disease | 389 (13.4%) | 81 (22.0%) | 106 (19.9%) | 21 (21.4%) | 12 (15.6%) | 749 (20.0%) | 74 (15.3%) | 37 (11.3%) |
| Liver disease | 97/2,893 (3.4%) | 11 (3.0%) | 27 (5.1%) | 2 (2.0%) | 16 (20.8%) | 162/3,744 (4.3%) | 13/484 (2.7%) | 17 (5.2%) |
| Cancer | 97/2,893 (3.4%) | 22 (6.0%) | 26 (4.9%) | 2 (2.0%) | 8 (10.4%) | 349/3,744 (9.3%) | 19/484 (3.9%) | 16 (4.9%) |
| Charlson Comorbidity Index | 2.0 (1.0 - 4.0) | 4.0 (1.0 - 6.0) | 4.0 (2.0 - 6.0) | 2.0 (1.0 - 5.0) | 4.0 (2.0 - 6.0) | 4.0 (3.0 - 7.0) | 3.0 (2.0 - 5.0) | 3.0 (1.0 - 5.0) |
| **Presenting Vital Signs** |  |  |  |  |  |  |  |  |
| Temperature ≥ 38°C | 655/2,604 (25.2%) | 75/313 (24.0%) | 120/497 (24.1%) | 15/90 (16.7%) | 15/71 (21.1%) | 700/3,410 (20.5%) | 78/429 (18.2%) | 60/295 (20.3%) |
| Oxygen saturation < 94% | 1,327 (45.8%) | 118 (32.1%) | 225 (42.2%) | 37 (37.8%) | 29 (37.7%) | 1,496 (39.9%) | 191 (39.4%) | 139 (42.6%) |
| Respiration rate > 24 breaths/min | 1,072/2,851 (37.6%) | 97/358 (27.1%) | 206/525 (39.2%) | 28/96 (29.2%) | 26/75 (34.7%) | 949/3,681 (25.8%) | 154/476 (32.4%) | 103/322 (32.0%) |
| **Baseline Clinical Labs** |  |  |  |  |  |  |  |  |
| White blood cell > 12 x10<sup>9</sup>/L | 454/2,827 (16.1%) | 50/353 (14.2%) | 68/525 (13.0%) | 17/97 (17.5%) | 16/75 (21.3%) | 426/3,656 (11.7%) | 49/473 (10.4%) | 39/312 (12.5%) |
| Lymphocyte < 1 x10<sup>9</sup>/L | 1,548/2,666 (58.1%) | 151/322 (46.9%) | 322/500 (64.4%) | 54/90 (60.0%) | 41/69 (59.4%) | 2,077/3,432 (60.5%) | 271/448 (60.5%) | 170/293 (58.0%) |
| Platelet, < 150,000 x10<sup>9</sup>/L | 466/2,825 (16.5%) | 66/353 (18.7%) | 111/525 (21.1%) | 20/97 (20.6%) | 12/75 (16.0%) | 882/3,654 (24.1%) | 112/473 (23.7%) | 54/312 (17.3%) |
| AST<sup>a</sup> > 40 U/L | 1,387/2,616 (53.0%) | 153/323 (47.4%) | 328/491 (66.8%) | 51/91 (56.0%) | 39/74 (52.7%) | 1,487/3,424 (43.4%) | 206/443 (46.5%) | 158/293 (53.9%) |
| ALT<sup>b</sup> > 40 U/L | 1,026/2,612 (39.3%) | 97/317 (30.6%) | 206/488 (42.2%) | 33/90 (36.7%) | 22/74 (29.7%) | 967/3,418 (28.3%) | 161/444 (36.3%) | 122/289 (42.2%) |
| BUN<sup>c</sup> > 20 mg/dL | 897/2,813 (31.9%) | 166/350 (47.4%) | 238/524 (45.4%) | 45/94 (47.9%) | 23/76 (30.3%) | 1,703/3,662 (46.5%) | 181/471 (38.4%) | 124/314 (39.5%) |
| Creatinine > 1.5 mg/dL | 412/2,816 (14.6%) | 97/351 (27.6%) | 96/525 (18.3%) | 23/95 (24.2%) | 14/76 (18.4%) | 640/3,668 (17.4%) | 83/471 (17.6%) | 49/314 (15.6%) |
| Total bilirubin > 1.2 mg/dL | 121/2,327 (5.2%) | 19/318 (6.0%) | 25/481 (5.2%) | 14/90 (15.6%) | 15/72 (20.8%) | 179/3,140 (5.7%) | 14/423 (3.3%) | 20/273 (7.3%) |
| Sodium < 130 mmol/L | 220/2,812 (7.8%) | 10/350 (2.9%) | 53/524 (10.1%) | 12/94 (12.8%) | 12/76 (15.8%) | 193/3,662 (5.3%) | 28/471 (5.9%) | 20/314 (6.4%) |
| **Baseline Clinical Status<sup>d</sup>** |  |  |  |  |  |  |  |  |
| WHO Score 3 | 1,506/2,891 (52.1%) | 225/366 (61.5%) | 255 (47.8%) | 52 (53.1%) | 50 (64.9%) | 2,134/3,747 (57.0%) | 254/483 (52.6%) | 188 (57.7%) |
| WHO Score 4 | 1,065/2,891 (36.8%) | 106/366 (29.0%) | 224 (42.0%) | 37 (37.8%) | 16 (20.8%) | 1,361/3,747 (36.3%) | 185/483 (38.3%) | 109 (33.4%) |
| WHO Score 5 | 228/2,891 (7.9%) | 22/366 (6.0%) | 37 (6.9%) | 6 (6.1%) | 9 (11.7%) | 185/3,747 (4.9%) | 31/483 (6.4%) | 16 (4.9%) |
| WHO Score 6 | 49/2,891 (1.7%) | 10/366 (2.7%) | 6 (1.1%) | 2 (2.0%) | 0 (0.0%) | 36/3,747 (1.0%) | 8/483 (1.7%) | 6 (1.8%) |
| WHO Score 7 | 43/2,891 (1.5%) | 3/366 (0.8%) | 11 (2.1%) | 1 (1.0%) | 2 (2.6%) | 31/3,747 (0.8%) | 5/483 (1.0%) | 7 (2.1%) |
| **Clinical Course** |  |  |  |  |  |  |  |  |
| ICU admissions | 930 (32.1%) | 105 (28.5%) | 201 (37.7%) | 33 (33.7%) | 25 (32.5%) | 1,144 (30.5%) | 192 (39.6%) | 107 (32.8%) |
| Mechanical ventilation | 537 (18.5%) | 58 (15.8%) | 92 (17.3%) | 16 (16.3%) | 12 (15.6%) | 417 (11.1%) | 79 (16.3%) | 55 (16.9%) |
| Hospital mortality | 408 (14.1%) | 52 (14.1%) | 80 (15.0%) | 13 (13.3%) | 16 (20.8%) | 568 (15.1%) | 69 (14.2%) | 40 (12.3%) |
| Discharged alive | 2,462 (85.0%) | 315 (85.6%) | 446 (83.7%) | 83 (84.7%) | 60 (77.9%) | 3,169 (84.5%) | 412 (84.9%) | 283 (86.8%) |
| Continued hospitalization | 28 (1.0%) | 1 (0.3%) | 7 (1.3%) | 2 (2.0%) | 1 (1.3%) | 14 (0.4%) | 4 (0.8%) | 3 (0.9%) |

<sup>a</sup> Aspartate transaminase
<sup>b</sup> Alanine transaminase
<sup>c</sup> Blood urea nitrogen
<sup>d</sup> Measured on the WHO Clinical Progression Scale, as defined as: WHO Score 3 - hospitalized, no oxygen therapy; WHO Score 4 - hospitalized, oxygen by mask or nasal prongs; WHO Score 5 - hospitalized, non-invasive ventilation or high-flow oxygen; WHO Score 6 - hospitalized, intubation and mechanical ventilation; WHO Score 7 - hospitalized, ventilation + additional organ support

On admission, a higher percentage of Hispanic patients (11.1%) than White patients (6.7%) had a score of five or above on the WHO Clinical Progression Scale, and a higher percentage of Hispanic patients than White patients were febrile, had low oxygen saturation, and had high respiration rates. Over the course of hospitalization, a higher percentage of Hispanic patients (18.5%) than White patients (11.1%) needed mechanical ventilation.

### Characteristics of SARS-CoV-2 Infected Patients Over Time

Throughout the pandemic, rates of positive test, hospitalization, and in-hospital mortality per 100,000 patients were on average higher for Black, Hispanic, NH/PI, and AI/AN patients than Asian and White patients (**Figure 1**). Hispanics patients, in particular, had the highest rates, especially during the June-July and November-December resurgence of COVID-19. The mean age of SARS-CoV-2 infected patients generally decreased over time while the mean age of COVID-19 hospitalized patients and patients who experienced in-hospital mortality remained relatively consistent.

**Figure 1.**
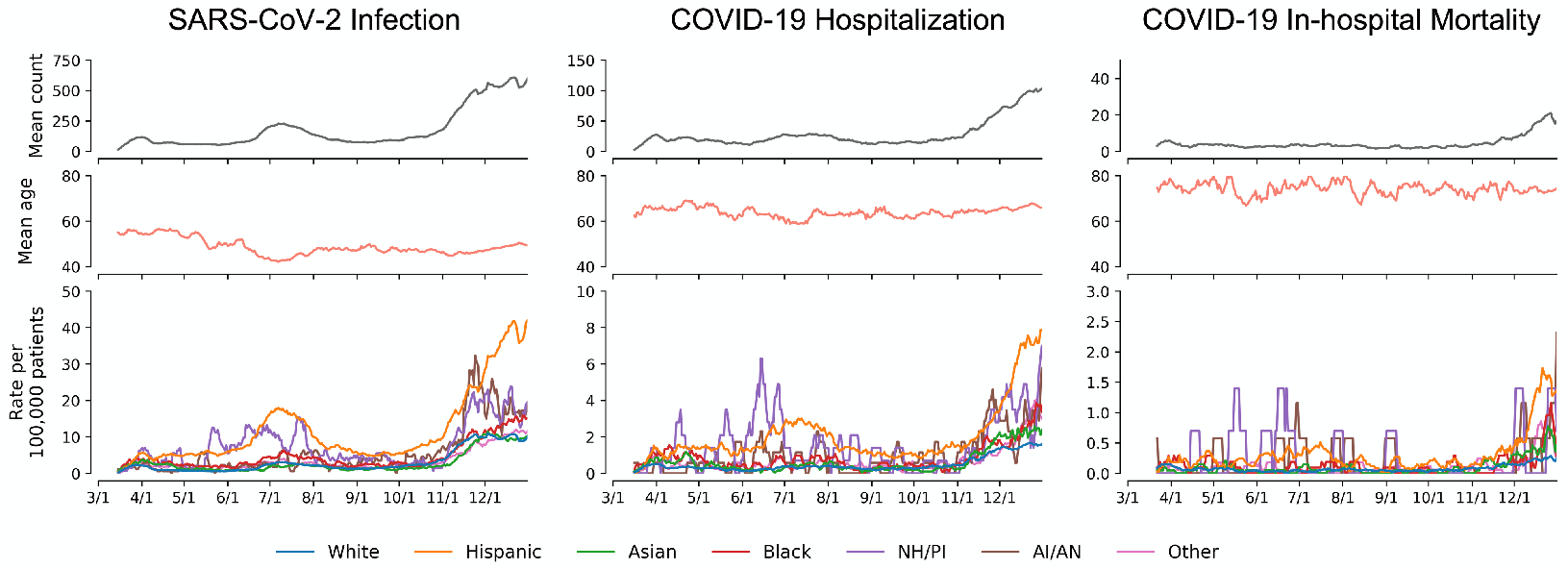
Characteristics of COVID-19 patients over time (rolling 7-day average). Left column represents patients who have tested positive for SARS-CoV-2 infection; middle column represents patients who have been hospitalized for COVID-19; and right column represents COVID-19 hospitalized patients who experienced in-hospital death. The first row of each column represents the rolling 7-day mean count of patients for the event; the second row represents the rolling 7-day mean age of patients; and the third row represents the 7-day mean rate of event per 100,000 patients. Rate per 100,000 patients were calculated out of total patients under care since 2019 for each race/ethnicity.

### Comparison with State-level SARS-CoV-2 Cases, COVID-19 Hospitalization, and Death

The distribution of race/ethnicity among patients in this study generally reflected state-level distributions for COVID-19 outcomes (**Figure S1-3**), including for mortality in which state-level data captured both in-hospital and out-of-hospital deaths. High proportions of Hispanics were consistently observed in both the state-wide data and the health system. The differences in racial/ethnic distributions between the health system population and the catchment population were consistent or smaller than the differences observed at the state-level (**Figure S4-5**).

### Factors Associated with SARS-CoV-2 Infection

Minority populations including Hispanic, Black, Asian, NH/PI and AI/AN had increased odds of SARS-CoV-2 infection compared to Whites in unadjusted and adjusted analysis (**Table 4**). In adjusted multivariate analysis, increased odds of positive SARS-CoV-2 results were independently associated with Hispanic (OR [95% CI]: 3.09 [2.99-3.18]), NH/PI (2.23 [2.01-2.48]), Black (1.35 [1.28 - 1.43]), Asian (1.31 [1.25 - 1.38]), and Other (1.69 [1.6 - 1.78]) race/ethnicity with White patients as the reference category. Increasing age and age-squared, male sex, overweight, obesity (all categories), Medicaid insurance, lack of insurance, and residence in a neighborhood with higher percentage limited English proficient individuals were also independently associated with increased odds of infection. Higher CCI score, Medicare insurance, and higher median income, however, were associated with lower odds of positive SARS-CoV-2 infection.

**Table 4.** Factors associated with SARS-CoV-2 infection, COVID-19 hospitalization, and COVID-19-related in-hospital mortality.

| Variable | Odds Ratio for Testing Positive |  | Odds ratio for Hospitalization |  | Odds ratio for In-Hospital Mortality |  |
| --- | --- | --- | --- | --- | --- | --- |
|  | Unadjusted | Multivariate | Unadjusted | Multivariate | Unadjusted | Multivariate |
| White | (ref) | (ref) | (ref) | (ref) | (ref) | (ref) |
| Hispanic | 4.21 (4.12 - 4.31) | 3.09 (2.99 - 3.18) | 0.85 (0.8 - 0.9) | 1.31 (1.22 - 1.42) | 0.89 (0.77 - 1.03) | 1.41 (1.15 - 1.71) |
| Black | 1.65 (1.57 - 1.73) | 1.35 (1.28 - 1.43) | 0.98 (0.87 - 1.1) | 1.18 (1.02 - 1.36) | 0.89 (0.65 - 1.21) | 1.05 (0.73 - 1.52) |
| Asian | 1.32 (1.26 - 1.38) | 1.31 (1.25 - 1.38) | 1.36 (1.23 - 1.52) | 1.62 (1.43 - 1.84) | 0.97 (0.75 - 1.25) | 0.93 (0.69 - 1.25) |
| NH/PI | 2.55 (2.31 - 2.82) | 2.23 (2.01 - 2.48) | 1.4 (1.11 - 1.77) | 2.01 (1.55 - 2.61) | 0.85 (0.47 - 1.54) | 1.17 (0.6 - 2.28) |
| AI/AN | 1.51 (1.36 - 1.67) | 1.33 (1.2 - 1.48) | 1.3 (1.01 - 1.67) | 1.56 (1.17 - 2.06) | 1.53 (0.87 - 2.68) | 1.92 (0.96 - 3.81) |
| Other | 1.92 (1.83 - 2.01) | 1.69 (1.6 - 1.78) | 1.12 (1 - 1.25) | 1.32 (1.16 - 1.5) | 0.88 (0.67 - 1.15) | 1.09 (0.8 - 1.49) |
| Unknown | 1.6 (1.55 - 1.65) | 1.49 (1.41 - 1.56) | 0.32 (0.28 - 0.36) | 0.65 (0.57 - 0.74) | 0.79 (0.56 - 1.12) | 1.01 (0.69 - 1.49) |
| Age (years) | 0.83 (0.82 - 0.83) | 1.12 (1.1 - 1.14) | 2.93 (2.85 - 3.01) | 2.32 (2.21 - 2.44) | 2.13 (1.97 - 2.29) | 5.37 (2.74 - 10.56) |
| Age-squared | 1.03 (1.02 - 1.04) | 1.13 (1.12 - 1.15) | 1.63 (1.59 - 1.66) | 0.86 (0.83 - 0.88) | 1.93 (1.81 - 2.06) | 0.42 (0.23 - 0.75) |
| Female | (ref) | (ref) | (ref) | (ref) | (ref) | (ref) |
| Male | 1.21 (1.19 - 1.23) | 1.27 (1.25 - 1.29) | 1.43 (1.36 - 1.5) | 1.6 (1.51 - 1.69) | 1.23 (1.09 - 1.39) | 1.1 (0.95 - 1.27) |
| CCI | 0.94 (0.94 - 0.95) | 0.97 (0.96 - 0.98) | 1.42 (1.4 - 1.43) | 1.15 (1.13 - 1.17) | 1.19 (1.17 - 1.21) | 1.07 (1.04 - 1.1) |
| Hypertension | 0.85 (0.83 - 0.87) | 0.97 (0.95 - 1) | 3.29 (3.12 - 3.46) | 1.12 (1.05 - 1.19) | 1.19 (1.05 - 1.34) | 0.72 (0.62 - 0.84) |
| Underweight | 0.92 (0.85 - 0.99) | 0.94 (0.87 - 1.03) | 2.2 (1.87 - 2.58) | 1.43 (1.18 - 1.74) | 1.1 (0.79 - 1.52) | 0.85 (0.59 - 1.21) |
| Normal | (ref) | (ref) | (ref) | (ref) | (ref) | (ref) |
| Overweight | 1.26 (1.22 - 1.3) | 1.15 (1.12 - 1.19) | 0.88 (0.83 - 0.94) | 0.89 (0.83 - 0.96) | 0.9 (0.77 - 1.05) | 1.04 (0.87 - 1.25) |
| Class 1 Obesity | 1.38 (1.25 - 1.52) | 1.21 (1.16 - 1.25) | 0.93 (0.87 - 1) | 1.06 (0.97 - 1.15) | 0.77 (0.65 - 0.93) | 1.05 (0.85 - 1.3) |
| Class 2 Obesity | 1.38 (1.18 - 1.62) | 1.24 (1.19 - 1.3) | 0.99 (0.91 - 1.08) | 1.32 (1.2 - 1.47) | 0.73 (0.58 - 0.92) | 1.14 (0.86 - 1.49) |
| Class 3 Obesity | 1.41 (1.15 - 1.74) | 1.25 (1.2 - 1.31) | 1.29 (1.17 - 1.41) | 1.99 (1.79 - 2.21) | 0.82 (0.65 - 1.04) | 1.56 (1.17 - 2.06) |
| Commercial Insurance | (ref) | (ref) | (ref) | (ref) | (ref) | (ref) |
| Medicaid | 1.49 (1.46 - 1.52) | 1.14 (1.11 - 1.17) | 3.82 (3.58 - 4.07) | 2.84 (2.64 - 3.04) | 2.87 (2.27 - 3.63) | 1.63 (1.25 - 2.12) |
| Medicare | 0.6 (0.59 - 0.62) | 0.59 (0.57 - 0.61) | 8.38 (7.79 - 9.02) | 2.05 (1.87 - 2.24) | 3.86 (3.03 - 4.92) | 1.48 (1.11 - 1.96) |
| Uninsured/Self-pay | 4.58 (4.27 - 4.91) | 2.85 (2.66 - 3.07) | 1.36 (1.11 - 1.67) | 1.57 (1.27 - 1.94) | 1.63 (0.82 - 3.24) | 1.27 (0.59 - 2.72) |
| Median income (log) | 0.72 (0.71 - 0.72) | 0.84 (0.83 - 0.85) | 0.96 (0.94 - 0.98) | 1.06 (1.03 - 1.1) | 0.98 (0.93 - 1.05) | 0.97 (0.89 - 1.05) |
| % Crowded housing | 1.39 (1.38 - 1.4) | 1.01 (0.99 - 1.03) | 1.06 (1.03 - 1.08) | 0.92 (0.87 - 0.98) | 1.06 (1 - 1.12) | 1.03 (0.87 - 1.21) |
| % Minority | 1.47 (1.46 - 1.49) | 0.99 (0.97 - 1.01) | 1.07 (1.03 - 1.1) | 0.92 (0.86 - 0.98) | 1.07 (1.01 - 1.14) | 1.07 (0.92 - 1.25) |
| % Limited English | 1.44 (1.43 - 1.45) | 1.13 (1.11 - 1.15) | 1.17 (1.13 - 1.2) | 1.33 (1.24 - 1.43) | 1 (0.93 - 1.08) | 0.96 (0.81 - 1.14) |
| WHO Score 3 | - | - | - | - | (ref) | (ref) |
| WHO Score 4 | - | - | - | - | 1.94 (1.68 - 2.22) | 1.79 (1.54 - 2.08) |
| WHO Score 5 | - | - | - | - | 7.52 (6.15 - 9.19) | 6.5 (5.17 - 8.17) |
| WHO Score 6 | - | - | - | - | 6.8 (4.62 - 10) | 5.32 (3.4 - 8.3) |
| WHO Score 7 | - | - | - | - | 9.65 (6.45 - 14.42) | 6.57 (4.15 - 10.38) |
| WBC > 12 x10<sup>9</sup>/L | - | - | - | - | 2.19 (1.91 - 2.52) | 1.67 (1.41 - 1.98) |
| Lymphocyte < 1 x10<sup>9</sup>/L | - | - | - | - | 1.5 (1.31 - 1.71) | 1.26 (1.08 - 1.47) |
| Platelet, < 150,000 x10<sup>9</sup>/L | - | - | - | - | 1.57 (1.37 - 1.81) | 1.32 (1.13 - 1.55) |
| AST > 40 U/L | - | - | - | - | 1.73 (1.53 - 1.96) | 1.66 (1.44 - 1.92) |
| BUN > 20 mg/dL | - | - | - | - | 4.18 (3.66 - 4.76) | 1.95 (1.66 - 2.31) |
| Creatinine > 1.5 mg/dL | - | - | - | - | 2.97 (2.59 - 3.4) | 1.38 (1.16 - 1.65) |
| Total bilirubin > 1.2 mg/dL | - | - | - | - | 1.91 (1.45 - 2.53) | 1.45 (1.00 - 2.11) |
| Sodium < 130 mmol/L | - | - | - | - | 1.33 (1.06 - 1.67) | 1.25 (0.96 - 1.61) |

### Factors Associated with Hospitalization for COVID-19

Minority races/ethnicities were also associated with increased odds for COVID-19 hospitalization (**Table 4**). In multivariate analysis, the odds ratio for COVID-19 hospitalization was highest among NH/PI patients (2.01 [1.55 - 2.61]), followed by Asian (1.62 [1.43 - 1.84]), AI/AN (1.56 [1.17 - 2.06]), Other (1.32 [1.16 - 1.5]), Hispanic (1.31 [1.22 - 1.42]), and Black (1.18 [1.02 - 1.36]) patients. Increasing age, male sex, public insurance, no insurance, higher CCI score, being underweight, having class 2 or 3 obesity, having hypertension, and residence in a neighborhood with higher rates of limited English proficiency were also independently associated with increased odds of hospitalization.

### Factors Associated with Hospital Mortality in Admissions for COVID-19

In adjusted multivariate analysis, Hispanic race/ethnicity was significantly associated with increased odds of in-hospital mortality (1.41 [1.15 - 1.71]). Other minority races/ethnicities, however, were not significantly associated (**Table 4**). Hospital mortality was also independently associated with age; class 3 obesity; public insurance; higher score on the WHO Clinical Progression Scale and CCI; and high white blood cell count, low lymphocyte, low platelet count, high AST, high blood urea nitrogen, high creatinine, and high bilirubin. Interaction analysis of race/ethnicity and age further identified disproportionately increased odds of hospital mortality among Hispanic patients as age increased (1.30 [1.04 - 1.62]; P_interaction_=0.021; **Figure S6**).

## Discussion

This study examined the characteristics and clinical outcomes of 629,953 patients tested for SARS-CoV-2 across California, Oregon, and Washington. Overall, we highlight how characteristics of SARS-CoV-2 infected patients vary by race/ethnicity and show differential associations of COVID-19 hospitalizations, morbidity, and mortality across race/ethnic sub-populations. While patients of minority race/ethnicity represented 27.8% of patients tested for SARS-CoV-2, they constituted 50.1% of SARS-CoV-2 infected patients and 54.3% of COVID-19 hospitalized patients. Hispanic patients in particular represented 34.3% of SARS-CoV-2 infected and 35.3% of hospitalized patients despite making up only 13.4% of tested patients, a pattern consistent with state-level statistics. Race/ethnicity was associated with SARS-CoV-2 infection and COVID-19 hospitalization, with increased odds of both outcomes highest among Hispanics and NH/PI patients. Hispanic race/ethnicity was also significantly associated with increased odds of hospital mortality. These findings showcase the disproportionate burden of COVID-19 born by Hispanics in these western US states, despite being younger age and having generally fewer comorbidities than White patients.

Understanding racial and ethnic disparities in COVID-19 cases and outcomes is important for understanding the nature of the disease and guiding public health prevention efforts and medical interventions. This study comprehensively detailed and compared the clinical and epidemiological characteristics of COVID-19 across all major races/ethnicities, highlighting in particular Asian, NH/PI, and AI/AN patients who have been underrepresented in COVID-19 literature to date. In doing so, we show that these populations, particularly NH/PI patients, have similar or higher odds of SARS-CoV-2 infection and hospitalization as Black and Hispanic patients. Concomitantly, we show that factors associated with COVID-19 severity vary across race/ethnicity. These findings demonstrate the need to further focus resources on addressing COVID-19 in all minority communities through efforts such as culturally-appropriate public health messaging, data collection, improving access to testing, and possibly actively intervene earlier in the disease course.[19]

The significant associations of minority races/ethnicities with SARS-CoV-2 infection and COVID-19 hospitalization builds on previous analyses of Black and Hispanic patients.[20–23] However, unlike previous studies, we also found a significant association between Hispanic race/ethnicity and hospital mortality and a significant interaction between Hispanic race/ethnicity and age that signifies the relationship between Hispanic race/ethnicity and hospital mortality is moderated by age. The clinical characteristics of Hispanic patients at hospital admission suggest that they are presenting with more severe illness than White patients, as Hispanic patients are more likely to be febrile, have low oxygen saturation, and high respiration rates. The higher percentage of Hispanic patients presented with WHO Clinical Progression Scale scores of five or higher reflects a need for high-flow supplemental oxygen or mechanical ventilation. These clinical findings indicate a delay in seeking care among Hispanic patients. While this study cannot identify the causes behind the observed associations, certain social, structural or biologic determinants of health have been suggested.[3] Social and structural determinants could include occupation risk and limited access to healthcare and testing.[3] Additional studies are needed to identify the causal factors driving disparities in COVID-19.

Racial/ethnic disparities continue to persist as the pandemic endures. Our temporal analysis revealed that resurgences of COVID-19 particularly burdened minority patients. As vaccines and treatments against COVID-19 are studied, meaningful representation of diverse populations in clinical trials, particularly of those disproportionately affected by COVID-19, is necessary to ensure their safety and efficacy.[24] Such representation can also help build public trust in the studied vaccine or treatment. Distribution of vaccines should also take into consideration racial equity, as it can help mitigate the disproportionate impact of the virus and prevent widening health disparities.[25] Unfortunately, minority populations face various barriers to vaccination, including lack of access to healthcare, distrust of the healthcare system, communication barriers, and misinformation. Thus, additional resources should be placed on reaching high-risk populations, building trust, and reducing barriers to vaccination among minorities.

While the large size of this study’s diverse cohort and its multi-state distribution are strengths of this study, there are limitations. This study was limited to a single health system and certain catchment areas within California, Oregon, and Washington. Thus, the results may be less generalizable to other regions. In particular, the racial/ethnicity composition of COVID-19 patients in our study’s may not reflect those in other states and at the national level. For example, Black patients are underrepresented in our study when compared to national-level statistics. At the same time, electronic health records are subject to the quality, consistency, and completeness of entry by providers. Some patients may have also received care at other institutions and therefore certain outcomes or characteristics may be underreported. Our use of ICD-10-CM code may not capture diagnoses that are not billable or represented by a given code. Despite these limitations, however, our results highlight how the impact of COVID-19 vary across race/ethnicity in a large geographical area.

## Data Availability

Data used in this study is archived within PSJH in a HIPAA-secure location to facilitate verification of study conclusions.

## Author Contributions

Concept and design: CLD, SAK, RTR, GD, JDG, and JJH

Acquisition, analysis, or interpretation of data: CLD, SAK, RTR, GD, JDG, JJH, and ATM Drafting of the manuscript: CLD

Critical revision of the manuscript for important intellectual content: All authors

Statistical analysis: CLD and SAK

Obtained funding: JRH, JDG, JJH, and ATM

Supervision: JJH and ATM

CLD, SAK, and RTR had full access to all of the data in the study and take responsibility for the integrity of the data and the accuracy of the data analysis.

## Acknowledgement

We thank the caregivers in the PSJH system for their tireless efforts in caring for patients with COVID-19. We also thank Mark Premo, Lindsay Mico, Jennifer Jones, Andrey Dubovoy, Vivek Tomer, and the PSJH Health Intelligence team for their support and management of the electronic health record data.

