## Supplemental Materials for "Characteristics and Factors Associated with COVID-19 Infection, Hospitalization, and Mortality Across Race and Ethnicity"

**Table S1. Characteristics of patients tested for SARS-CoV-2 infection**

|  | **Hispanic No. (%)** | **Black No. (%)** | **Asian No. (%)** | **NH/PI No. (%)** | **AI/AN No. (%)** | **White No. (%)** | **Other No. (%)** | **Unknown No. (%)** |
| --- | --- | --- | --- | --- | --- | --- | --- | --- |
| **Socio-demographics** |  |  |  |  |  |  |  |  |
| Total no. of patients | 76,300 | 21,709 | 30,736 | 3,460 | 5,201 | 411,852 | 21,040 | 59,655 |
| COVID-19 positive | 16,836 (22.1%) | 2,066 (9.5%) | 2,456 (8.0%) | 468 (13.5%) | 434 (8.3%) | 24,468 (5.9%) | 2,353 (11.2%) | 5,564 (9.3%) |
| Age (years) | 44.1 ± 17.8 | 48.2 ± 18.2 | 51.1 ± 18.8 | 45.5 ± 17.3 | 47.3 ± 17.9 | 53.9 ± 19.4 | 49.7 ± 19.1 | 47.2 ± 18.4 |
| Sex |  |  |  |  |  |  |  |  |
| Female | 45,190 (59.2%) | 12,240 (56.4%) | 19,246 (62.6%) | 2,042 (59.0%) | 3,203 (61.6%) | 234,072 (56.8%) | 11,561 (54.9%) | 31,692 (53.1%) |
| Male | 31,110 (40.8%) | 9,469 (43.6%) | 11,490 (37.4%) | 1,418 (41.0%) | 1,998 (38.4%) | 177,780 (43.2%) | 9,479 (45.1%) | 27,963 (46.9%) |
| Insurance |  |  |  |  |  |  |  |  |
| Commercial | 29,509/73,826 (40.0%) | 7,392/21,259 (34.8%) | 17,473/30,211 (57.8%) | 1,599/3,348 (47.8%) | 1,376/5,135 (26.8%) | 180,018/407,457 (44.2%) | 9,205/20,582 (44.7%) | 33,728/54,230 (62.2%) |
| Medicaid | 37,013/73,826 (50.1%) | 10,582/21,259 (49.8%) | 7,729/30,211 (25.6%) | 1,307/3,348 (39.0%) | 3,036/5,135 (59.1%) | 106,762/407,457 (26.2%) | 8,150/20,582 (39.6%) | 9,968/54,230 (18.4%) |
| Medicare | 5,553/73,826 (7.5%) | 3,061/21,259 (14.4%) | 4,888/30,211 (16.2%) | 396/3,348 (11.8%) | 707/5,135 (13.8%) | 119,487/407,457 (29.3%) | 3,052/20,582 (14.8%) | 8,897/54,230 (16.4%) |
| Uninsured/Self-pay | 1,675/73,826 (2.3%) | 216/21,259 (1.0%) | 115/30,211 (0.4%) | 44/3,348 (1.3%) | 16/5,135 (0.3%) | 1,142/407,457 (0.3%) | 167/20,582 (0.8%) | 1,626/54,230 (3.0%) |
| Other insurance | 76/73,826 (0.1%) | 8/21,259 (0.0%) | 6/30,211 (0.0%) | 2/3,348 (0.1%) | 0 (0.0%) | 48/407,457 (0.0%) | 8/20,582 (0.0%) | 11/54,230 (0.0%) |
| Neighborhood-level |  |  |  |  |  |  |  |  |
| Median income | 66,741.0 ± 19,675.1 | 68,785.7 ± 21,156.2 | 83,126.2 ± 26,430.2 | 70,830.7 ± 20,242.4 | 65,979.8 ± 19,763.8 | 74,940.5 ± 25,277.4 | 77,143.0 ± 25,446.5 | 80,143.2 ± 28,126.2 |
| % Crowded housing | 9.0 ± 7.4 | 6.1 ± 5.5 | 4.9 ± 4.3 | 4.7 ± 4.5 | 3.5 ± 2.7 | 3.3 ± 3.1 | 5.1 ± 4.8 | 4.6 ± 5.0 |
| % Minority | 54.5 ± 27.1 | 49.4 ± 25.5 | 44.3 ± 19.9 | 39.0 ± 21.8 | 29.6 ± 18.1 | 28.3 ± 16.3 | 40.8 ± 20.8 | 37.0 ± 21.8 |
| % Limited English | 17.3 ± 11.1 | 13.2 ± 9.0 | 12.7 ± 8.3 | 10.5 ± 8.2 | 6.0 ± 5.9 | 7.2 ± 6.7 | 12.2 ± 9.1 | 9.8 ± 8.6 |
| **Comorbidities** |  |  |  |  |  |  |  |  |
| Hypertension | 12,795 (16.8%) | 6,787 (31.3%) | 7,314 (23.8%) | 922 (26.6%) | 1,317 (25.3%) | 107,194 (26.0%) | 4,333 (20.6%) | 6,295 (10.6%) |
| Diabetes | 8,140 (10.7%) | 3,045 (14.0%) | 3,726 (12.1%) | 590 (17.1%) | 719 (13.8%) | 38,738 (9.4%) | 2,002 (9.5%) | 2,492 (4.2%) |
| BMI |  |  |  |  |  |  |  |  |
| Underweight | 851/68,886 (1.2%) | 439/19,814 (2.2%) | 1,105/27,289 (4.0%) | 41/3,062 (1.3%) | 76/4,776 (1.6%) | 8,086/371,297 (2.2%) | 455/18,974 (2.4%) | 666/34,697 (1.9%) |
| Normal | 14,047/68,886 (20.4%) | 4,739/19,814 (23.9%) | 12,817/27,289 (47.0%) | 677/3,062 (22.1%) | 1,076/4,776 (22.5%) | 108,964/371,297 (29.3%) | 5,702/18,974 (30.1%) | 11,012/34,697 (31.7%) |
| Overweight | 22,422/68,886 (32.5%) | 5,800/19,814 (29.3%) | 8,988/27,289 (32.9%) | 813/3,062 (26.6%) | 1,246/4,776 (26.1%) | 116,941/371,297 (31.5%) | 6,386/18,974 (33.7%) | 11,509/34,697 (33.2%) |
| Class 1 Obesity | 17,183/68,886 (24.9%) | 4,212/19,814 (21.3%) | 3,116/27,289 (11.4%) | 716/3,062 (23.4%) | 1,051/4,776 (22.0%) | 72,419/371,297 (19.5%) | 3,635/18,974 (19.2%) | 6,501/34,697 (18.7%) |
| Class 2 Obesity | 8,339/68,886 (12.1%) | 2,337/19,814 (11.8%) | 862/27,289 (3.2%) | 397/3,062 (13.0%) | 689/4,776 (14.4%) | 35,768/371,297 (9.6%) | 1,573/18,974 (8.3%) | 2,949/34,697 (8.5%) |
| Class 3 Obesity | 6,044/68,886 (8.8%) | 2,287/19,814 (11.5%) | 401/27,289 (1.5%) | 418/3,062 (13.7%) | 638/4,776 (13.4%) | 29,119/371,297 (7.8%) | 1,223/18,974 (6.4%) | 2,060/34,697 (5.9%) |
| Chronic respiratory disease |  |  |  |  |  |  |  |  |
| Asthma | 4,170 (5.5%) | 1,906 (8.8%) | 1,591 (5.2%) | 283 (8.2%) | 543 (10.4%) | 29,078 (7.1%) | 1,224 (5.8%) | 2,013 (3.4%) |
| COPD | 960 (1.3%) | 965 (4.4%) | 495 (1.6%) | 94 (2.7%) | 338 (6.5%) | 21,060 (5.1%) | 578 (2.7%) | 823 (1.4%) |
| Cardiovascular disease |  |  |  |  |  |  |  |  |
| Coronary artery disease | 1,725 (2.3%) | 984 (4.5%) | 1,440 (4.7%) | 184 (5.3%) | 280 (5.4%) | 27,763 (6.7%) | 1,008 (4.8%) | 1,165 (2.0%) |
| Myocardial infarction | 1,030 (1.3%) | 581 (2.7%) | 686 (2.2%) | 97 (2.8%) | 120 (2.3%) | 10,473 (2.5%) | 460 (2.2%) | 532 (0.9%) |
| Congestive heart failure | 2,267 (3.0%) | 1,543 (7.1%) | 1,192 (3.9%) | 226 (6.5%) | 342 (6.6%) | 26,107 (6.3%) | 955 (4.5%) | 1,043 (1.7%) |
| Kidney disease | 3,472 (4.6%) | 2,012 (9.3%) | 1,749 (5.7%) | 337 (9.7%) | 351 (6.7%) | 25,039 (6.1%) | 1,007 (4.8%) | 1,187 (2.0%) |
| Liver disease | 2,748 (3.6%) | 698 (3.2%) | 1,373 (4.5%) | 108 (3.1%) | 333 (6.4%) | 13,081 (3.2%) | 631 (3.0%) | 800 (1.3%) |
| Cancer | 2,432 (3.2%) | 1,113 (5.1%) | 1,893 (6.2%) | 160 (4.6%) | 253 (4.9%) | 30,132 (7.3%) | 1,014 (4.8%) | 1,456 (2.4%) |
| Charlson Comorbidity Index | 0.0 (0.0 - 2.0) | 1.0 (0.0 - 3.0) | 1.0 (0.0 - 3.0) | 1.0 (0.0 - 3.0) | 1.0 (0.0 - 3.0) | 1.0 (0.0 - 4.0) | 1.0 (0.0 - 3.0) | 0.0 (0.0 - 1.0) |

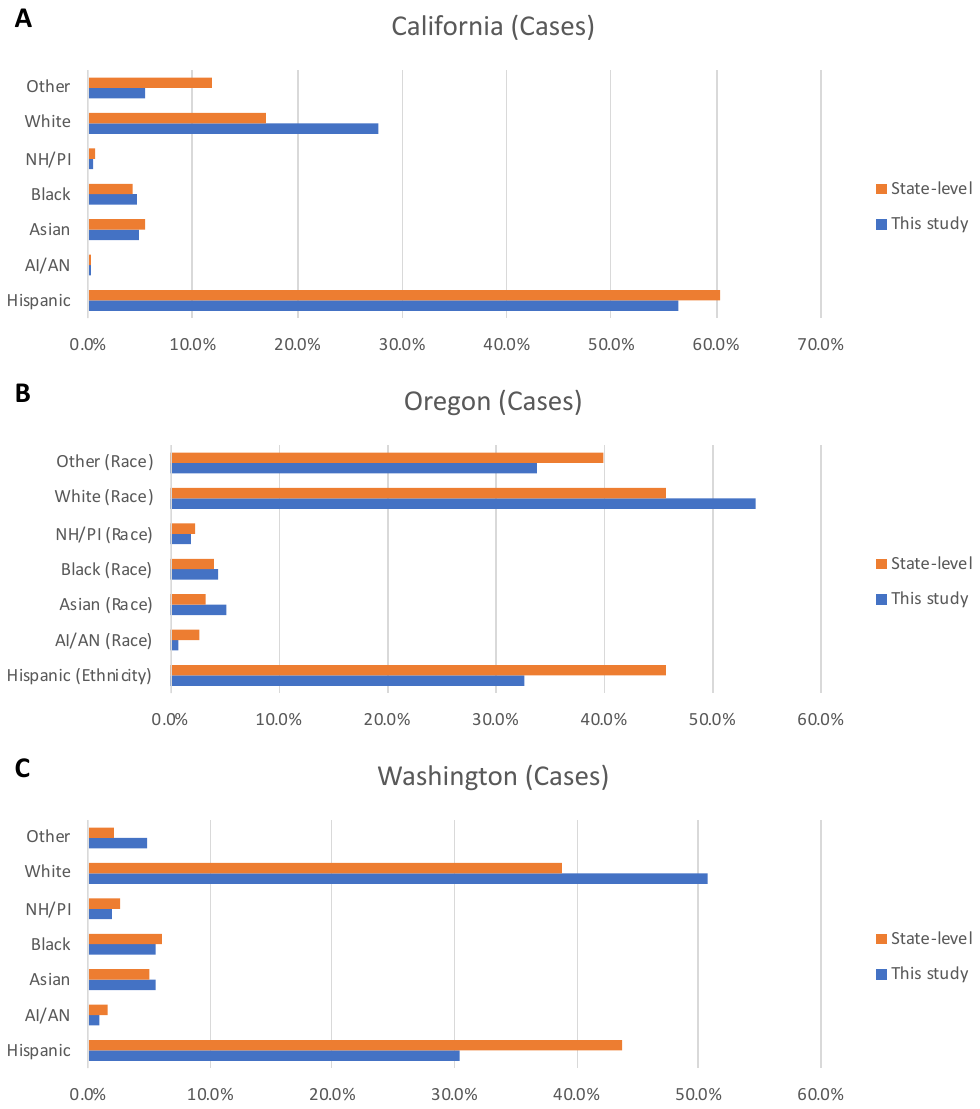

**Figure S1. Comparison of racial/ethnic distributions of SARS-CoV-2 infected patients with state-level distributions.** State-level data (orange bars) was derived from the COVID Racial Data Tracker project. Blue bars represent data from this study. Proportions are out of individuals with known race/ethnicity. While race and ethnicity are combined into one category for California (A) and Washington (C), they are separate categories for Oregon (B). Data from this study was therefore grouped to be consistent with state level data.

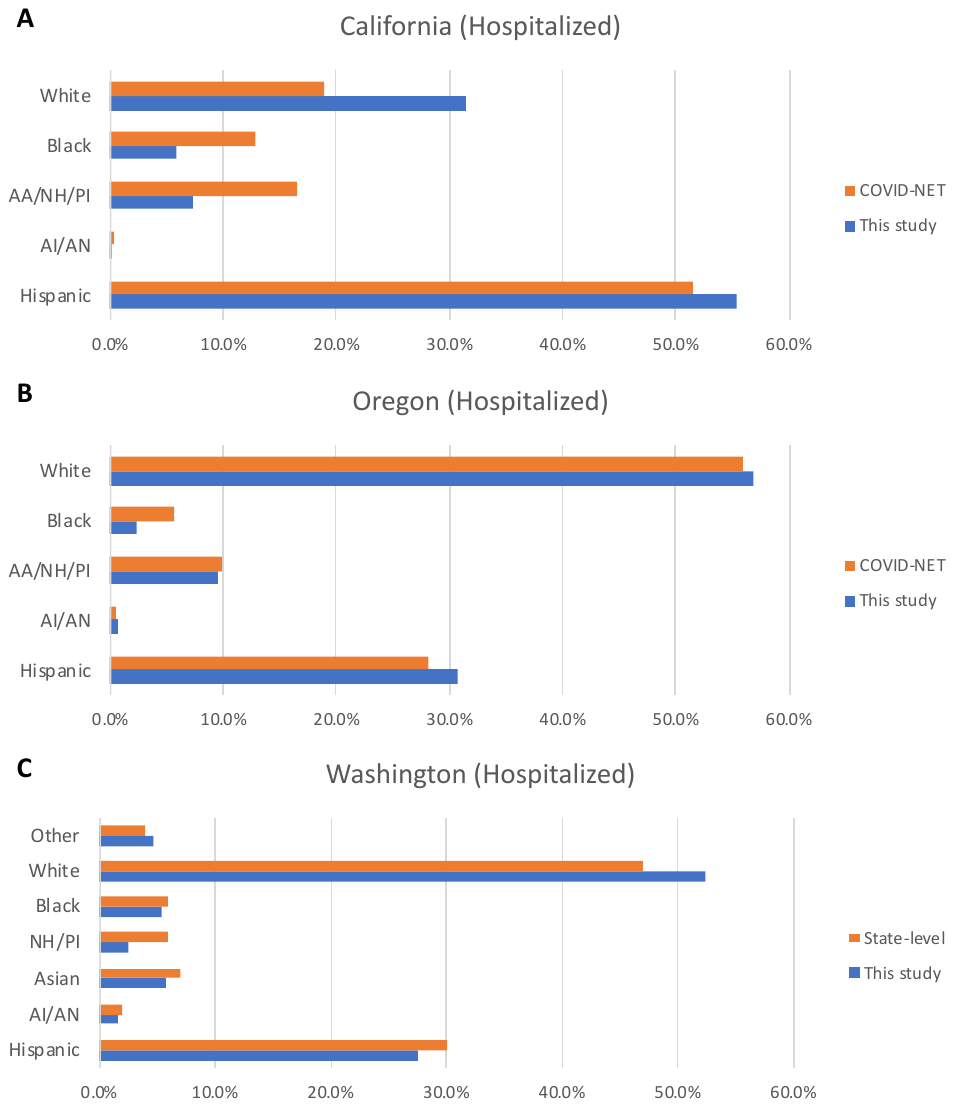

**Figure S2. Comparison of racial/ethnic distributions of COVID-19 hospitalization with state-level distributions.** State-level data (orange bars) was derived from the Center for Disease Control’s Coronavirus Disease 2019 (COVID-19)-Associated Hospitalization Surveillance Network data. Blue bars represent data from this study. Proportions are out of individuals with known race/ethnicity. For consistency, data from this study was grouped into racial/ethnic subgroups that are consistent with the state-level data.

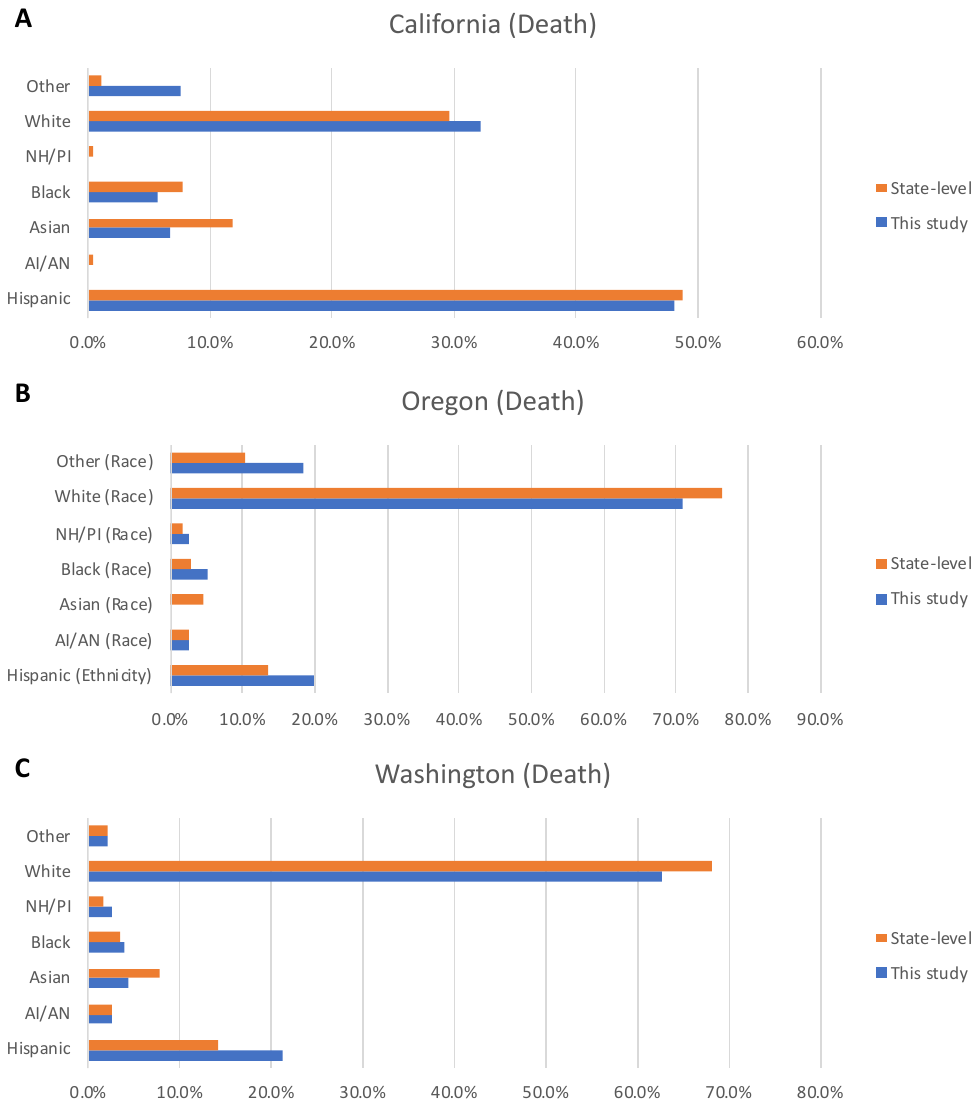

**Figure S3. Comparison of racial/ethnic distributions of COVID-19 deaths with state-level distributions.**

Death at the state-level include both in-hospital and out-of-hospital death while deaths in this study represented only in-hospital deaths. State-level data (orange bars) was derived from the COVID Racial Data Tracker project. Blue bars represent data from this study. Proportions are out of individuals with known race/ethnicity. While race and ethnicity are combined into one category for California (A) and Washington (C), they are separate categories for Oregon (B). For consistency, data from this study was grouped into racial/ethnic subgroups that are consistent with the state-level data.

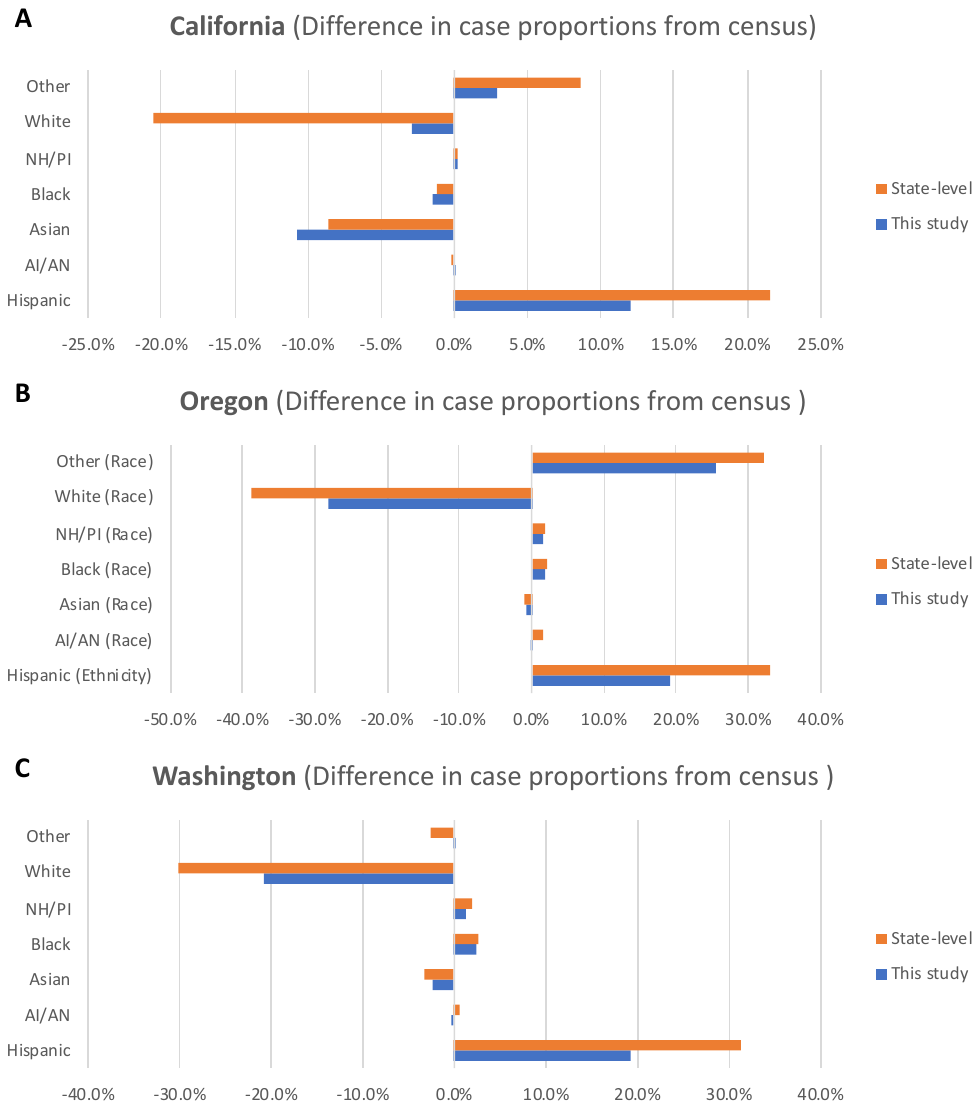

**Figure S4. Differences in racial/ethnic distributions of SARS-CoV-2 infected patients with population distributions.** Comparison of racial/ethnic distributions of COVID-19 cases with the underlying population served. Population served was calculated using census data. For state-level data (orange bars), state-wide census distributions were used. For data in this study (blue bars), the underlying population was weighted by the ZIP codes which had patients tested for COVID-19. For consistency, data from this study was grouped into racial/ethnic subgroups that are consistent with the state-level data.

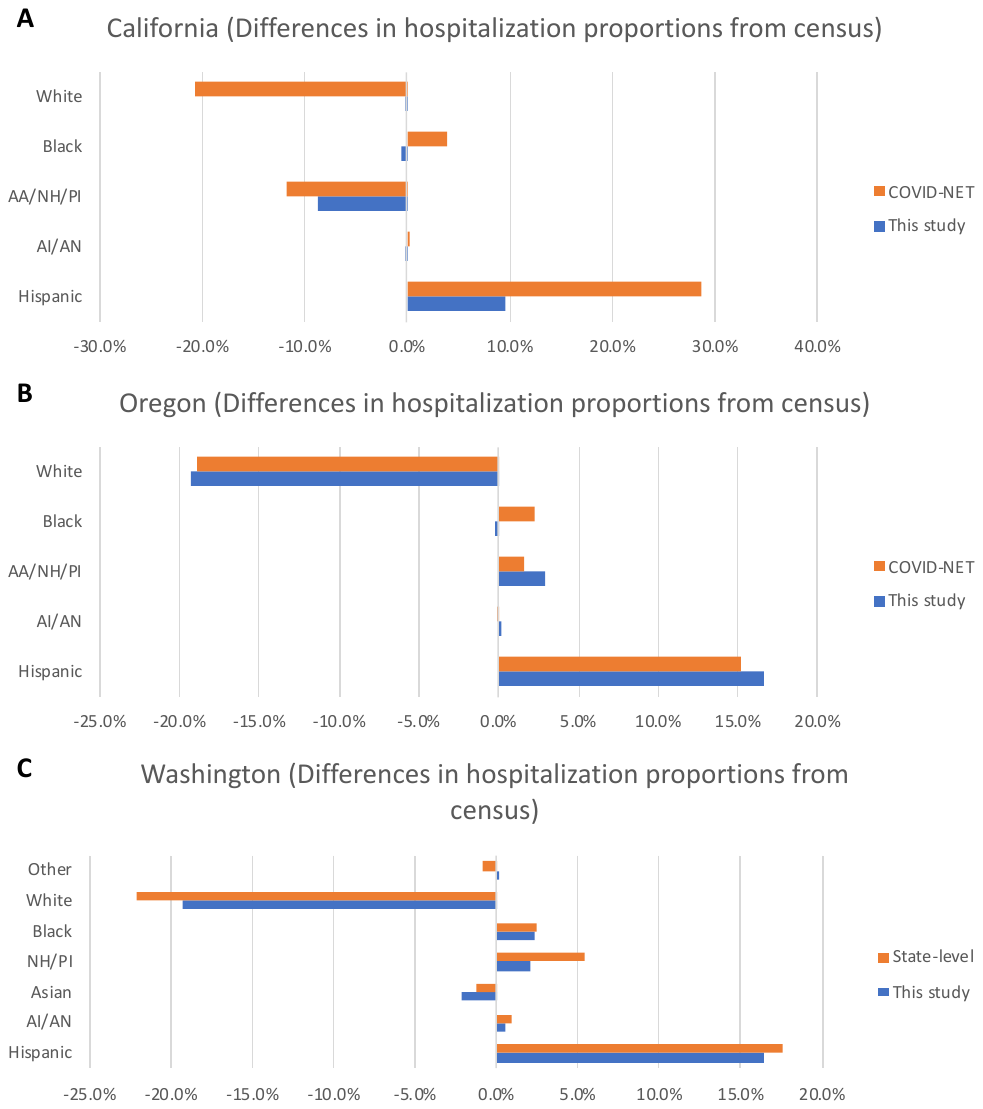

**Figure S5. Differences in racial/ethnic distributions of COVID-19 hospitalization with population distributions.** Comparison of racial/ethnic distributions of COVID-19 hospitalizations with the underlying population served. Population served was calculated using census data. For COVID-NET data (orange bars), populations were represented by the counties represented in the COVID-NET dataset. For Washington (C), state-wide census distributions were used. For data in this study (blue bars), the underlying population was weighted by the ZIP codes which had patients tested for COVID-19. For consistency, data from this study was grouped into racial/ethnic subgroups that are consistent with the state-level data.

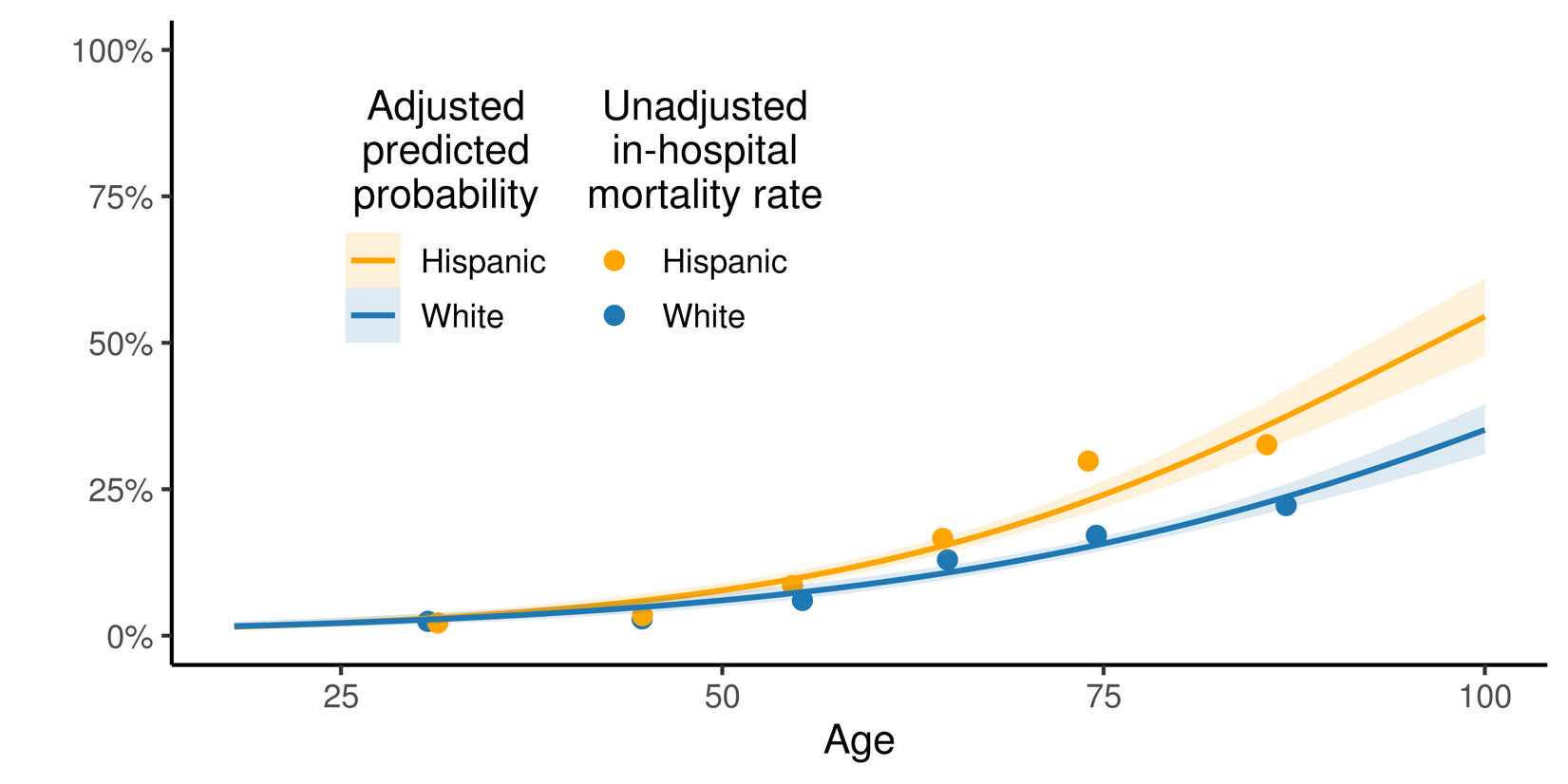

**Figure S6. Unadjusted and adjusted hospital mortality by race/ethnicity and age group.**

Orange points represents unadjusted hospital mortality rates for Hispanic patients and blue points represents rates for White patients. Rates were calculated by age group: 18-39, 40-49, 50-59, 60-69, 70-79, and 80+. Orange line represents adjusted predicted probabilities for hospital mortality for Hispanic patients and blue line represents predicted probabilities for White patients. Adjusted predicted probabilities were calculated with fully-adjusted multivariate logistic regression with an interaction-term between race/ethnicity and age.
